# The association between ABO blood group and SARS-CoV-2 infection: a meta-analysis

**DOI:** 10.1101/2020.07.17.20155986

**Authors:** Davide Golinelli, Erik Boetto, Elisa Maietti, Maria Pia Fantini

## Abstract

At present, existing evidence about the association between SARS-CoV-2 infection and ABO blood group polymorphism is preliminary and controversial. In this meta-analysis we investigate this association and determine SARS-CoV-2 positive individuals’ odds of having a specific blood group compared to controls.

We performed a systematic search on MEDLINE and LitCovid databases for studies published through July 15, 2020. Seven studies met inclusion criteria for meta-analysis, including a total of 13 subgroups of populations (7503 SARS-CoV-2 positive cases and 2962160 controls).

We analysed the odds of having each blood group among SARS-CoV-2 positive patients compared with controls. Random-effects models were used to obtain the overall pooled odds ratio (OR). Subgroup and sensitivity analyses were performed in order to explore the source of heterogeneity and results consistency.

The results of our meta-analysis indicate that SARS-CoV-2 positive individuals are more likely to have blood group A (pooled OR 1.23, 95%CI: 1.09-1.40) and less likely to have blood group O (pooled OR=0.77, 95%CI: 0.67-0.88).

Further studies are needed to investigate the mechanisms at the basis of this association, which may affect the kinetics of the pandemic according to the blood group distribution within the population.

## Introduction

Since the first outbreak of COVID-19 in Wuhan, China, researchers have extensively analysed the characteristics, clinical presentation and risk factors of individuals with SARS-CoV-2 infection.^1-4^ Among the risk and predisposing factors of many infectious diseases the ABO blood group polymorphism has been studied for many pathogens, starting from the pioneering studies by Helmbold and Vogel in the early ‘60s.^5^ Several researchers investigated the host susceptibility linked to the blood group or anti-histo-blood group antibodies for Coronaviridae viruses (e.g. SARS-CoV^6^) and other viral families like Retroviridae (e.g. HIV^7^) or Hepadnaviridae (e.g. HBV^8^).

In 2005, Cheng et al. reported that SARS-CoV infection susceptibility in a group of health care workers in Hong Kong, exposed to an index SARS patient, was influenced by the ABO blood group systems; in particular, compared with non-O blood group hospital staff, blood group O hospital staff had a lower chance of getting infected.^6^ On a further investigation, Guillon et al. found that either a monoclonal anti-A antibody or natural plasma anti-A specifically inhibited the SARS-CoV spike (S) protein/ACE2-dependent adhesion to ACE2-expressing cell lines.^9^ Therefore, ABO polymorphism could have contributed to substantially influence SARS-CoV susceptibility, affecting both the number of infected individuals and the kinetics of the 2002-2003 SARS-CoV epidemic.

Lu et al. reported a structural similarity between the receptor-binding domains of SARS-CoV and SARS-CoV-2;^10^ SARS-CoV and SARS-CoV-2 also use the same receptor, ACE2, for entry into target cells.^10,11^ This has led researchers to investigate whether the ABO blood group polymorphism is also associated with host susceptibility to SARS-CoV-2 infection. The hypotheses related to this association include the prevalence and distribution of specific genetic loci ^11,12^ translating proteins involved in a particular response to the infection (e.g. receptors or receptor binding proteins ^13,14^) - in terms of reduced or increased entrance capacity of the virus into the host cells -, the presence of a particular antibody population,^9^ and so on.

Despite the short time passed since the beginning of the pandemic, the speed at which science is moving has led to the publication of many studies suggesting the possible association between specific blood groups and SARS-CoV-2, mainly in the form of case-control studies.

However, whether a specific blood group is associated with an increased risk of SARS-CoV-2 infection, and the strength of this association, remains preliminary and controversial. Therefore, the primary objective of this study is to verify the presence and strength of the ABO blood group polymorphism association with SARS-CoV-2 infection through a meta-analysis of the available epidemiological data. Particularly, we aim to determine SARS-CoV-2 positive individuals’ odds of having a specific blood group compared to controls, characteristics which may affect the kinetics of the pandemic according to the blood group distribution within the population.

## Materials and methods

### Databases and search strategy

We systematically searched for studies published through July 15, 2020 without restrictions on publication date or language. We searched the MEDLINE and LitCovid databases using the following search terms, respectively: *(COVID-19 OR SARS-CoV-2) AND (“blood group” OR “ABO”)*; *”COVID-19” OR “SARS-CoV-2” AND “blood group” OR “ABO”*. The initial inquiry was conducted by one of us (E.B.) and independently verified by another (D.G.).

### Eligibility criteria

Eligibility was restricted to studies examining the association between SARS-CoV-2 infection and ABO blood groups. As for study design, no specific criteria were used for inclusion. However, given the research question and topic of our study, we mainly found case-control and case series studies.

Studies were included only if reporting adequate data on the control group (i.e. blood group distribution among not SARS-CoV-2 positive individuals). We also included reports, correspondence and letters to the editor, if reporting original data.

COVID-19 literature also includes a number of studies on the association between SARS-CoV-2 infection and ABO blood groups available on preprint servers (e.g. MedrXiv). To maintain the quality of the data as high as possible, we decided to include only articles indexed on MEDLINE and LitCovid.

### Selection process

Two of us (E.B. and D.G.) independently reviewed titles and abstracts of the retrieved studies. Based on the selection criteria, both reviewers independently identified eligible studies. Disagreements were resolved by discussion, with a prior agreement that any unsettled conflict would be determined by a third reviewer (MP.F.).

Data collection forms (Excel spreadsheet [Microsoft Corporation, Redmond, WA]) were used by both reviewers to extract the required data from eligible studies. Both reviewers were unblinded to the studies’ authors’ names, population sizes, journals of publication, and locations.

### Data extraction

We extracted data on the blood group distribution in the population of SARS-CoV-2 positive (SARS-CoV-2+) individuals and in control groups, and the related information (e.g. characteristics of cases and control population). Other data of interest included the following study information: first author name, location, setting where the study population was enrolled and date of recruitment, if available.

Some studies reported more than one group of cases along with a corresponding control population. We included in the analysis all the comparisons regarding different sub-groups of cases in order to avoid any overlapping. We identified each study using a numerical ID followed by an alphabetic letter to distinguish different case-control comparisons.

Primary outcome measures were Odds Ratios (ORs) for the association of testing positive for SARS-CoV-2 (infection confirmed by Polymerase Chain Reaction, PCR) and having a specific blood group (A, B, AB, or O).

### Study quality assessment

Two of us (E.B. and D.G.) independently used the Newcastle-Ottawa Scale (NOS) for assessing the individual quality of each study (Supplementary material). NOS is specifically used for nonrandomized studies and has been endorsed by the Cochrane collaboration.^15^ Both authors independently evaluated all studies. Disagreements were resolved by discussion, with a prior agreement that any unsettled conflict would be determined by a third author (MP.F.). We argue that in the pandemic phase the risk of publication bias is negligible because the interest in having evidence on the new pathogen overcomes the possibility of not divulging data. For this reason tests or correction for publication bias were not deemed necessary.

### Statistical analysis

In order to analyse the association between SARS-CoV-2 and blood group, we compared each blood group against the others and computed group specific ORs with 95% confidence interval (95%*CI*). An OR greater than 1 indicates that SARS-CoV-2 patients were more likely to have a specific blood group than controls. Conversely an odds ratio smaller than 1 indicates that SARS-CoV-2 patients are less likely to have a specific blood group than controls.

The Stata command *metan* was used to perform meta-analysis of ORs and to derive pooled estimates. A random-effects analysis was performed using the DerSimonian and Laird method, with the estimate of heterogeneity being taken from the inverse-variance.

Results were graphically displayed through the forest plot. In order to evaluate the influence of each study on the pooled OR, sensitivity analyses were conducted using the leave-one-out approach. Heterogeneity was assessed using Cochran’s Q test and *I^2^* statistic. *I^2^* > 50% was considered to denote substantial heterogeneity and in such cases the sources of heterogeneity were explored with subgroup analyses. The studies were grouped according to the type of control population (general, blood donors, hospitalized) and country (China, USA, other).

The significance level was set at p<0.05. All statistical analyses were performed using Stata version 16 (StataCorp, 2019. *Stata Statistical Software: Release 16*. College Station, TX).

## Results

### Study screening

The literature search of the MEDLINE and LitCovid databases yielded 55 records. After the removal of 12 duplicates, we screened titles and abstracts excluding 26 records for lack of adherence to the inclusion criteria. We reviewed the full text of the remaining 17 studies and assessed their reference lists for relevant publications. No additional relevant publications were found. Based on this review, we excluded ten full-text articles for not meeting our inclusion criteria. A total of seven studies were included in the qualitative synthesis and meta-analysis.^16-22^ The flow diagram of the systematic literature review can be found in Fig 1.

**Fig 1.**
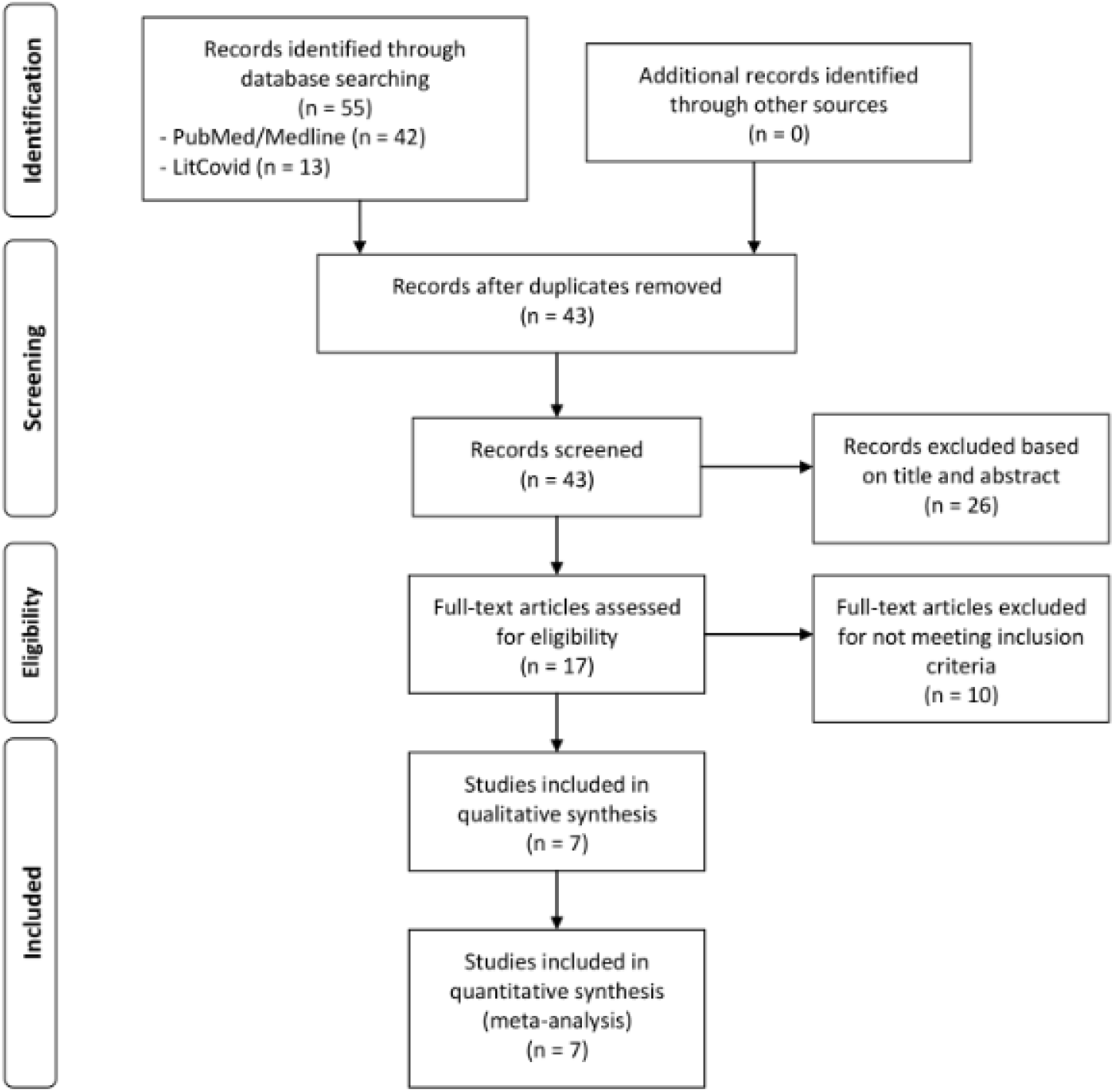
Flow Diagram of Systematic Literature Search for the Meta-analysis.

### Study quality assessment

The NOS version for case-control studies was used to address subject selection, study comparability and the outcome or exposure. NOS scores ranged from 4 to 5 points (8 being the highest possible score), with a mean of 4.7 and a median of 5. A summary of the evaluation is shown in Table S1 (Supplementary material).

### Meta-analysis

Our meta-analysis comprised a total of seven studies, which included 13 different subgroups of cases, each compared with a specific control group (for a total of 7503 cases and 2962160 controls), as reported in Table 1. Three studies comprising six different comparisons were conducted in the United States (USA) [ID:2,^17^ 6a-6d,^21^ 7^22^], two studies regarding four different comparisons in China [ID:1a-1c,^16^ 4^19^], one study with two comparisons in Europe [ID:5a(Italy),5b(Spain)^20^], one study with one comparison in Turkey [ID:3^18^]. Most studies reported data on SARS-CoV-2+ patients who were admitted to hospital. As for control groups, out of 13 comparisons, seven retrieved data on healthy individuals from blood donors databases [ID:3,5a,5b,6a,6b,6c,6d], four used the general population as control group deriving data from previous published studies or from institutional electronic health records [ID:1a,1b,1c,2], two recruited patients hospitalized previously than SARS-CoV-2 outbreak [ID:4,7].

**Table 1.** Characteristics of studies subgroups and cases/controls populations included in the meta-analysis.

| **Study** | **ID** | **Design and population** | **Cases characteristics, source and recruitment dates**<br>**(all confirmed SARS-CoV-2+)** | **Controls characteristics, source and recruitment dates** | **NOS score** |
| --- | --- | --- | --- | --- | --- |
| **Li et al.<sup>16</sup>** | **1a** | Case-control<br>Cases:1775<br>Controls:3694 | Patients from Jinyintan Hospital, Wuhan, Hubei province, China.<br>Recruitment: N/A | Normal population of Wuhan, Hubei province, China. from a previous study*.<br>Recruitment: N/A | 5 |
|  | **1b** | Case-control<br>Cases:113<br>Controls:3694 | Patients from Renmin Hospital of Wuhan University, Wuhan, Hubei province, China.<br>Recruitment: N/A | Normal population of Wuhan, Hubei province, China. from a previous study*.<br>Recruitment: N/A |  |
|  | **1c** | Case-control<br>Cases:265<br>Controls:3694 | Patients diagnosed who died or were discharged from Central Hospital of Wuhan, Wuhan, Hubei province, China.<br>Recruitment: N/A | Normal population of Wuhan, Hubei province, China. from a previous study*.<br>Recruitment: N/A |  |
| **Zietz et al.<sup>17</sup>** | **2** | Case-control<br>Cases:682<br>Controls:108860 | Patients from the electronic health record (EHR) system of New York Presbyterian/ Columbia University Irving Medical Center (NYP/CUIMC) hospital, New York, USA.<br>Recruitment: up to April 05, 2020. | Individuals recorded in the NYP/CUIMC EHR system, New York, USA, excluding results for any individuals later tested for COVID-19 (regardless of result).<br>Recruitment: May 2011 - June 2019. | 5 |
| **Göker et al.<sup>18</sup>** | **3** | Case-control<br>Cases:186<br>Controls:1881 | Patients who were followed at Hacettepe University School of Medicine Hospitals, Ankara, Turkey.<br>Recruitment: March 10, 2020 - May 05, 2020. | Healthy individuals who applied to the Hacettepe University Blood Bank, Ankara, Turkey.<br>Recruitment: March 01, 2011 - May 01, 2019. | 5 |
| **Wu et al.<sup>19</sup>** | **4** | Case-control<br>Cases:187<br>Controls:1991 | Patients hospitalized in or discharged from First Hospital of Changsha, Changsha, Hunan province, China.<br>Recruitment: January 20, 2020 - March 5, 2020. | Non-COVID-19 Han Chinese patients with determined blood types who were hospitalized in or discharged from First Hospital of Changsha, Changsha, Hunan province, China.<br>Recruitment: January 2019 - February 2020. | 4 |
| **Ellinghaus et al.<sup>20</sup>** | **5a** | Case-control<br>Cases:835<br>Controls:1255 | Patients with severe Covid-19, defined as hospitalization with respiratory failure, from: Fondazione IRCCS Cà Granda Ospedale Maggiore Policlinico, Milan (597 patients); Humanitas Clinical and Research Center, IRCCS, Milan (154 patients); UNIMIB (Università degli Studi di Milano-Bicocca) School of Medicine, San Gerardo Hospital, Monza (200 patients); all in Italy.<br>Recruitment: N/A | Randomly selected blood donors at Fondazione IRCCS Cà Granda Ospedale Maggiore Policlinico, Milan; healthy volunteers, blood donors, and outpatients of gastroenterology departments in Italy.<br>Recruitment: N/A | 5 |
|  | **5b** | Case-control<br>Cases:775<br>Controls:950 | Patients with severe Covid-19, defined as hospitalization with respiratory failure, from: Hospital Clinic and IDIBAPS (Institut de Investigacions Biomèdiques August Pi i Sunyer), Barcelona (56 patients); Hospital Universitario Vall d'Hebron, Barcelona (337 patients); Hospital Universitario Ramón y Cajal, Madrid (298 patients); Donostia University Hospital, San Sebastian (338 patients); all in Spain.<br>Recruitment: N/A | Healthy blood donors in San Sebastian, Spain.<br>Recruitment: N/A |  |
| **Leaf et al.<sup>21</sup>** | **6a** | Case-control<br>Cases:561<br>Controls:2215626 | White non-hispanic patients admitted to ICU, from the Study of the Treatment and Outcomes in critically ill Patients with COVID-19 (STOP-COVID, collecting data from 67 hospitals across the United States.<br>Recruitment: March 04, 2020 - April 11, 2020. | White non-hispanic blood donors in the United States, from a previous study**.<br>Recruitment: 1991 - 2000. | 5 |
|  | **6b** | Case-control<br>Cases:645<br>Controls:23605<br>0 | Black non-hispanic patients admitted to ICU, from the Study of the Treatment and Outcomes in critically ill Patients with COVID-19 (STOP-COVID), collecting data from 67 hospitals across the United States.<br>Recruitment: March 04, 2020 - April 11, 2020. | Black non-hispanic blood donors in the United States, from a previous study**. Recruitment: 1991 - 2000. |  |
|  | **6c** | Case-control<br>Cases:114<br>Controls:12678<br>0 | Asian non-hispanic patients admitted to ICU, from the Study of the Treatment and Outcomes in critically ill Patients with COVID-19 (STOP-COVID), collecting data from 67 hospitals across the United States.<br>Recruitment: March 04, 2020 - April 11, 2020. | Asian non-hispanic blood donors in the United States, from a previous study**. Recruitment: 1991 - 2000. |  |
|  | **6d** | Case-control<br>Cases:408<br>Controls:25923<br>3 | Hispanic patients admitted to ICU, from the Study of the Treatment and Outcomes in critically ill Patients with COVID-19 (STOP-COVID), collecting data from 67 hospitals across the United States.<br>Recruitment: March 04, 2020 - April 11, 2020. | Hispanic blood donors in the United States, from a previous study**. Recruitment: 1991 - 2000. |  |
| **Dzik et al.<sup>22</sup>** | **7** | Case-control<br>Cases:957<br>Controls:5840 | Patients from Massachusetts General Hospital (MGH, n=745) and Brigham and Women's Hospital (BWH, n=212), Boston, USA.<br>Recruitment: February 12, 2020 - May 13, 2020. | Randomly selected patients who were hospitalized at MGH and BWH, Boston, USA.<br>Recruitment: March, 2019 - April 2019. | **4** |

\* Xu P, Xiong Y, Cao K. Distribution of ABO and RhD blood group among Healthy Han population in Wuhan. J Clin Hematol (China). 2015(28):837.
\*\* Garratty G, Glynn SA, McEntire R, Retrovirus Epidemiology Donor S. ABO and Rh(D) phenotype frequencies of different racial/ethnic groups in the United States. Transfusion 2004;44:703-6.

Blood group A frequency varies from 27% to 51% in cases and from 26% to 41% in controls (Table 2). Blood group B varies from 7% to nearly 33% in both groups, blood group AB frequency ranges from 2.5% to 13% in SARS-CoV-2+ patients and from 2.5 to 10% in controls, while blood group O varies between 22% to 61% in cases and from 30% to 57% in controls. This large variability indicates that ABO distribution was not similar among studies neither in cases or in controls.

**Table 2.** Cases and controls blood group distribution for study subgroup.

| Study | ID and Country | Population | Blood group distribution* |  |  |  |
| --- | --- | --- | --- | --- | --- | --- |
|  |  |  | A (%) | B (%) | AB (%) | O (%) |
| **Li et al.<sup>16</sup>** | **1a**<br>China | Cases, n=1775 | 37.7% | 26.4% | 10.0% | 25.8% |
|  |  | Controls, n=3694 | 32.2% | 24.9% | 9.1% | 33.8% |
|  | **1b**<br>China | Cases, n=113 | 39.8% | 22.1% | 13.3% | 24.8% |
|  |  | Controls, n=3694 | 32.2% | 24.9% | 9.1% | 33.8% |
|  | **1c**<br>China | Cases, n=265 | 39.2% | 25.3% | 9.8% | 25.7% |
|  |  | Controls, n=3694 | 32.2% | 24.9% | 9.1% | 33.8% |
| **Zietz et al.<sup>17</sup>** | **2**<br>USA | Cases, n=682 | 34.2% | 17.0% | 3.1% | 45.7% |
|  |  | Controls, n=108860 | 32.7% | 14.9% | 4.2% | 48.1% |
| **Göker et al.<sup>18</sup>** | **3**<br>Turkey | Cases, n=186 | 57.0% | 10.8% | 7.5% | 24.7% |
|  |  | Controls, n=1881 | 38.1% | 14.7% | 10.0% | 37.2% |
| **Wu et al.<sup>19</sup>** | **4**<br>China | Cases, n=187 | 36.9% | 33.7% | 7.5% | 21.9% |
|  |  | Controls, n=1991 | 27.5% | 32.3% | 10.0% | 30.2% |
| **Ellinghaus et al.<sup>20</sup>** | **5a**<br>Italy | Cases, n=835 | 46.5% | 10.9% | 5.1% | 37.5% |
|  |  | Controls, n=1255 | 35.9% | 13.0% | 4.0% | 47.1% |
|  | **5b**<br>Spain | Cases, n=775 | 48.6% | 9.2% | 4.6% | 37.5% |
|  |  | Controls, n=950 | 41.9% | 6.8% | 2.6% | 48.6% |
| **Leaf et al.<sup>21</sup>** | **6a**<br>USA<br>White non-hispanic | Cases, n=561 | 45.1% | 11.4% | 5.7% | 37.8% |
|  |  | Controls, n=2215626 | 39.7% | 10.9% | 4.1% | 45.2% |
|  | **6b**<br>USA<br>Black non-hispanic | Cases, n=645 | 27.1% | 21.7% | 3.3% | 47.9% |
|  |  | Controls, n=236050 | 25.8% | 19.7% | 4.3% | 50.2% |
|  | **6c**<br>USA<br>Asian non-hispanic | Cases, n=114 | 28.1% | 32.5% | 9.6% | 29.8% |
|  |  | Controls, n=126780 | 27.8% | 25.4% | 7.1% | 39.8% |
|  | **6d**<br>USA<br>Hispanic | Cases, n=408 | 29.4% | 6.9% | 2.5% | 61.3% |
|  |  | Controls, n=259233 | 31.1% | 9.9% | 2.5% | 56.5% |
| **Dzik et al.<sup>22</sup>** | **7**<br>USA | Cases, n=957 | 32.5% | 14.6% | 4.3% | 48.6% |
|  |  | Controls, n=5840 | 36.4% | 13.0% | 4.0% | 46.6% |

\*Percentages may not add up to 100.0% because of rounding.

### Blood group A association with SARS-CoV-2

The association of SARS-CoV-2 with blood group A was significant with a pooled OR of 1.23 (95%CI: 1.09-1.40), although the random-effect meta-analysis revealed a large heterogeneity among studies, *I^2^*=77.3%. The forest plot is shown in Fig 2.

**Fig 2.**
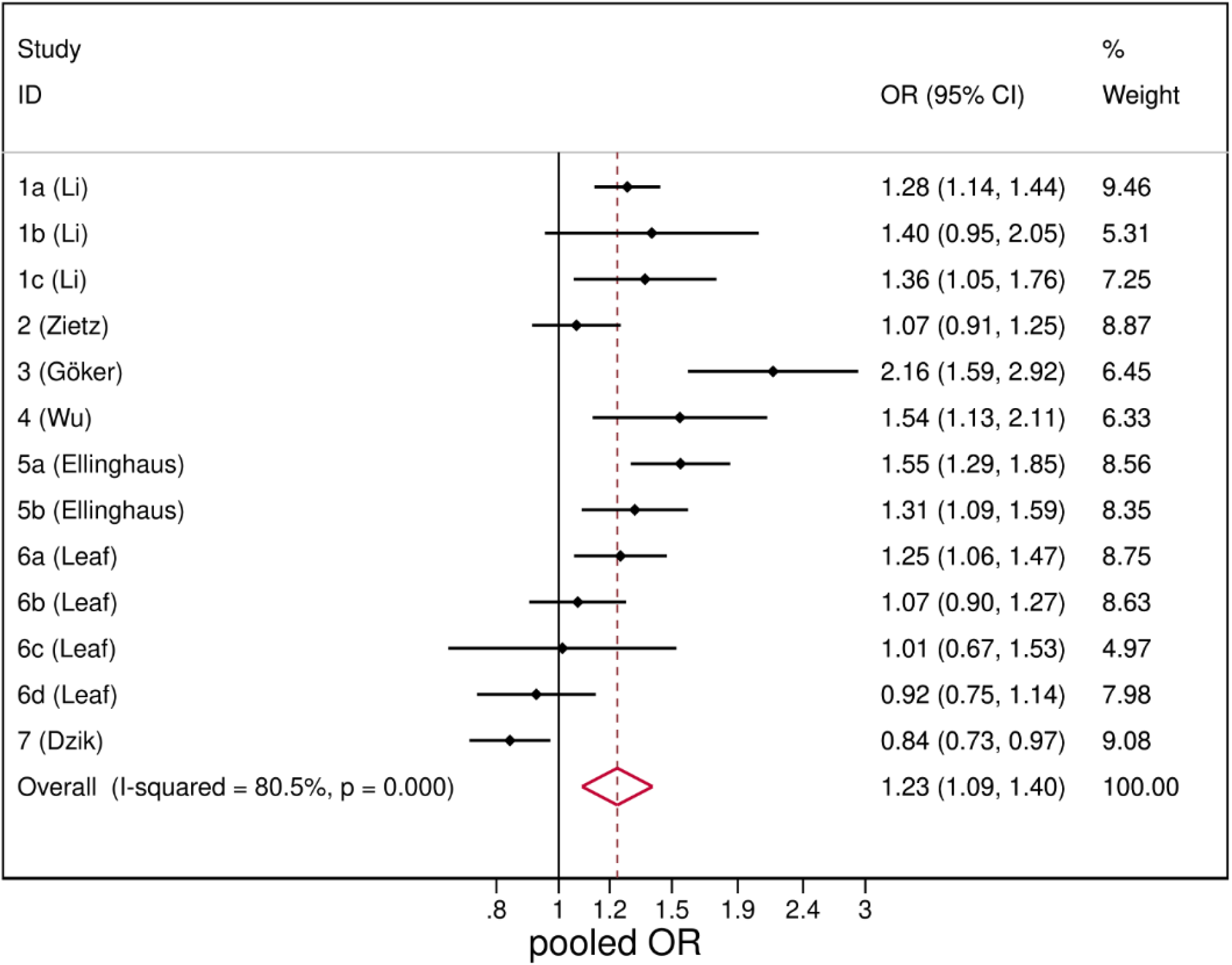
Forest plot from random effects analysis: OR of being blood group A in SARS-CoV-2+ group versus control group.

In sensitivity analysis the combined estimates indicate that none of the studies significantly affected the pooled estimate (Table S2, Supplementary material).

The type of control population did not account significantly for the observed heterogeneity; however we observed a greater homogeneity among studies that employed the general population (*I^2^*=34.4%) as compared to the others (Fig S1, Supplementary material). In the same way we observed homogeneity among studies conducted in China (*I^2^*=0%) (Fig S2, Supplementary material). In all these subgroups the pooled OR resulted concordant with the overall estimate indicating a positive association between blood group A and SARS-CoV-2+.

### Blood group B association with SARS-CoV-2

SARS-CoV-2 infection was unrelated with blood group B, with a pooled OR of 1.05 (95%CI: 0.96 -1.15) (Fig 3). The heterogeneity resulted quite low, I^2^=36.4% and the results from sensitivity analysis were consistent with the overall analysis (Table S2 Supplementary material).

**Fig 3.**
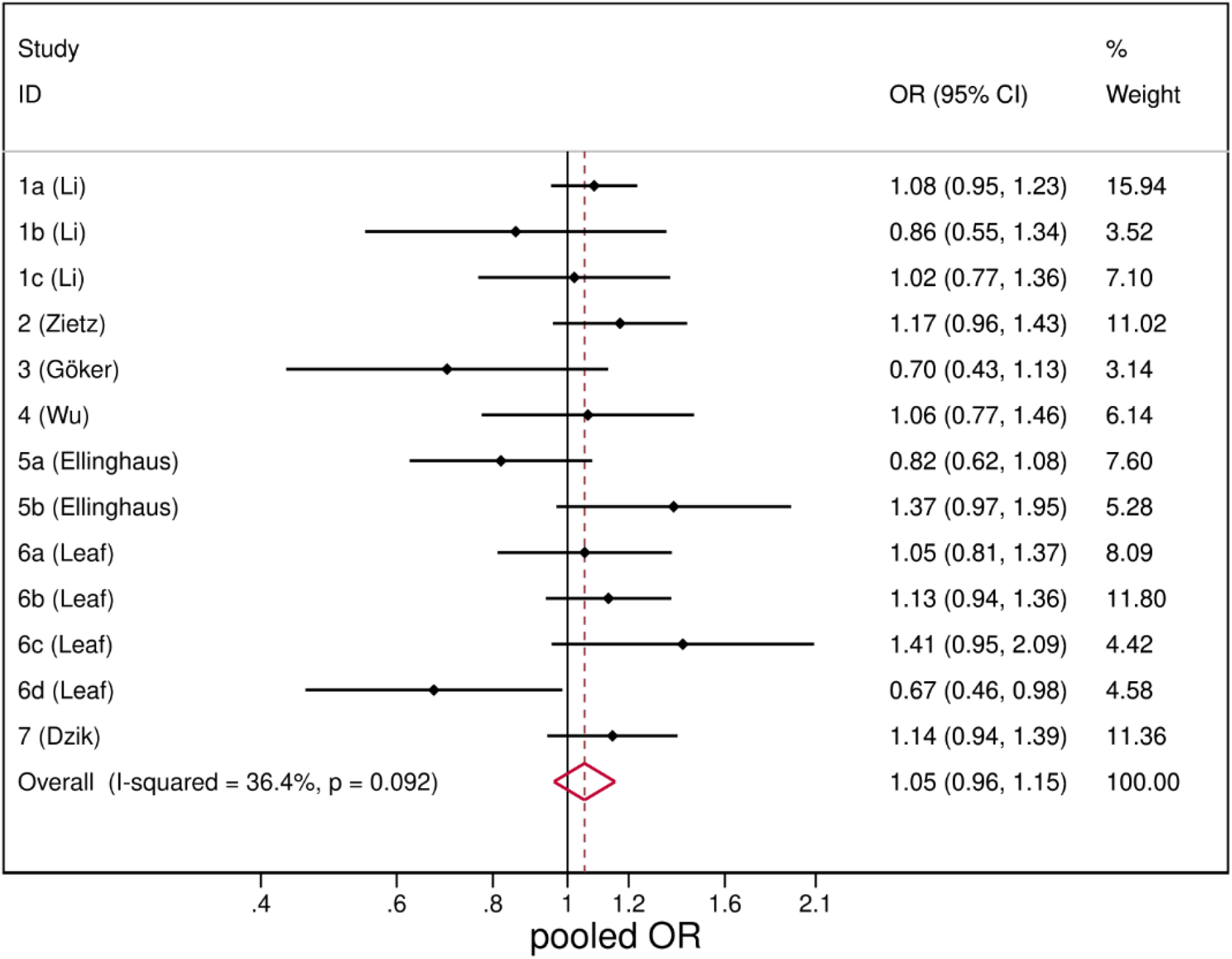
Forest plot from random effects analysis: OR of being blood group B in SARS-CoV-2+ group versus control group.

### Blood group AB association with SARS-CoV-2

SARS-CoV-2 infection was unrelated with blood group B (Fig 4), with a pooled OR of 1.09 (95%CI: 0.94 - 1.26). The heterogeneity was low (I^2^=35.9%) and results from sensitivity analysis were consistent with the main analysis (Table S2, Supplementary material).

**Fig 4.**
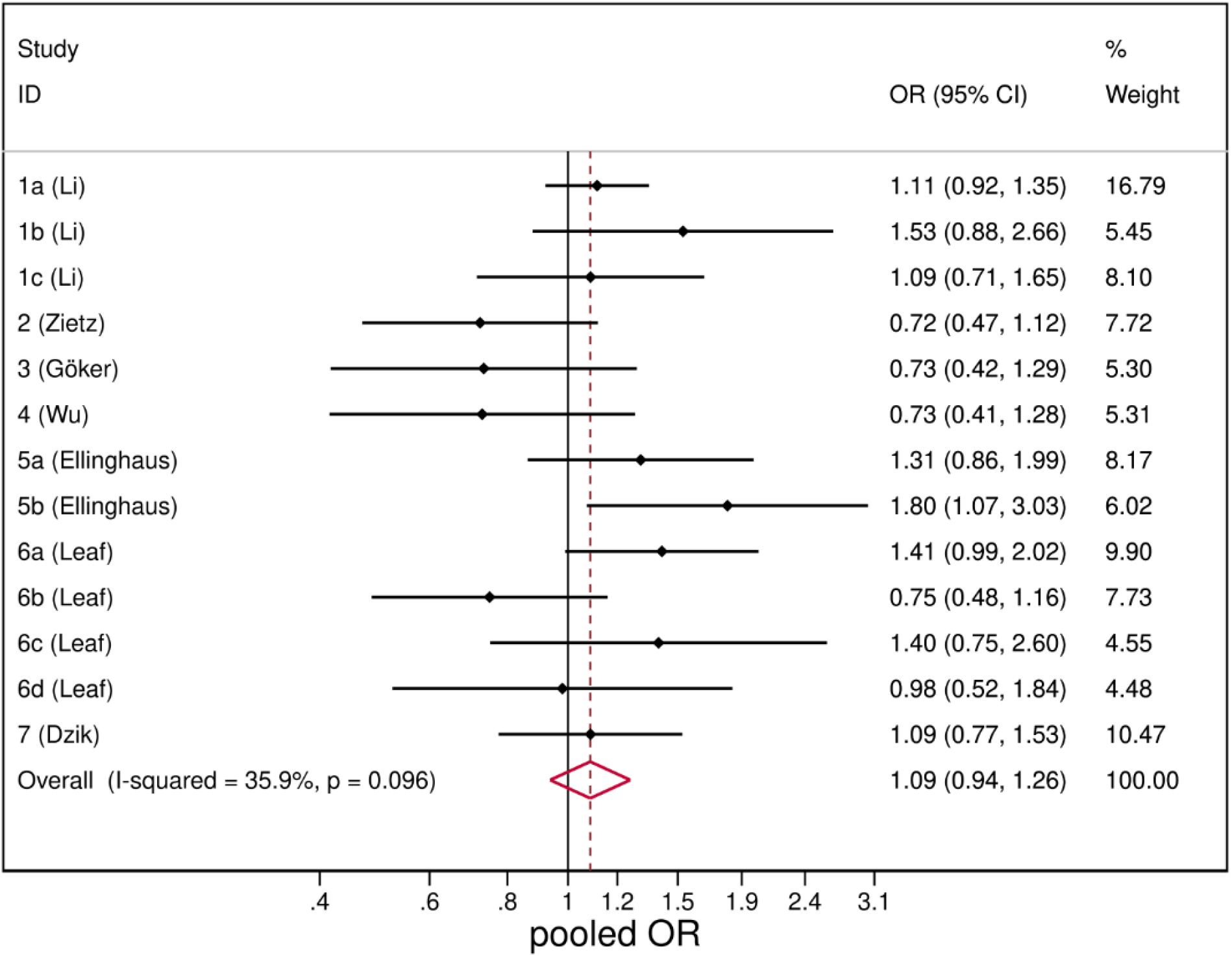
Forest plot from random effects analysis: OR of being blood group AB in SARS-CoV-2+ group versus control group.

### Blood group O association with SARS-CoV-2

Patients with SARS-CoV-2 infection were less likely to have blood group O (pooled OR=0.77, 95%CI: 0.67-0.88). However there was a very large heterogeneity *I^2^*=82% (Fig 5).

**Fig 5.**
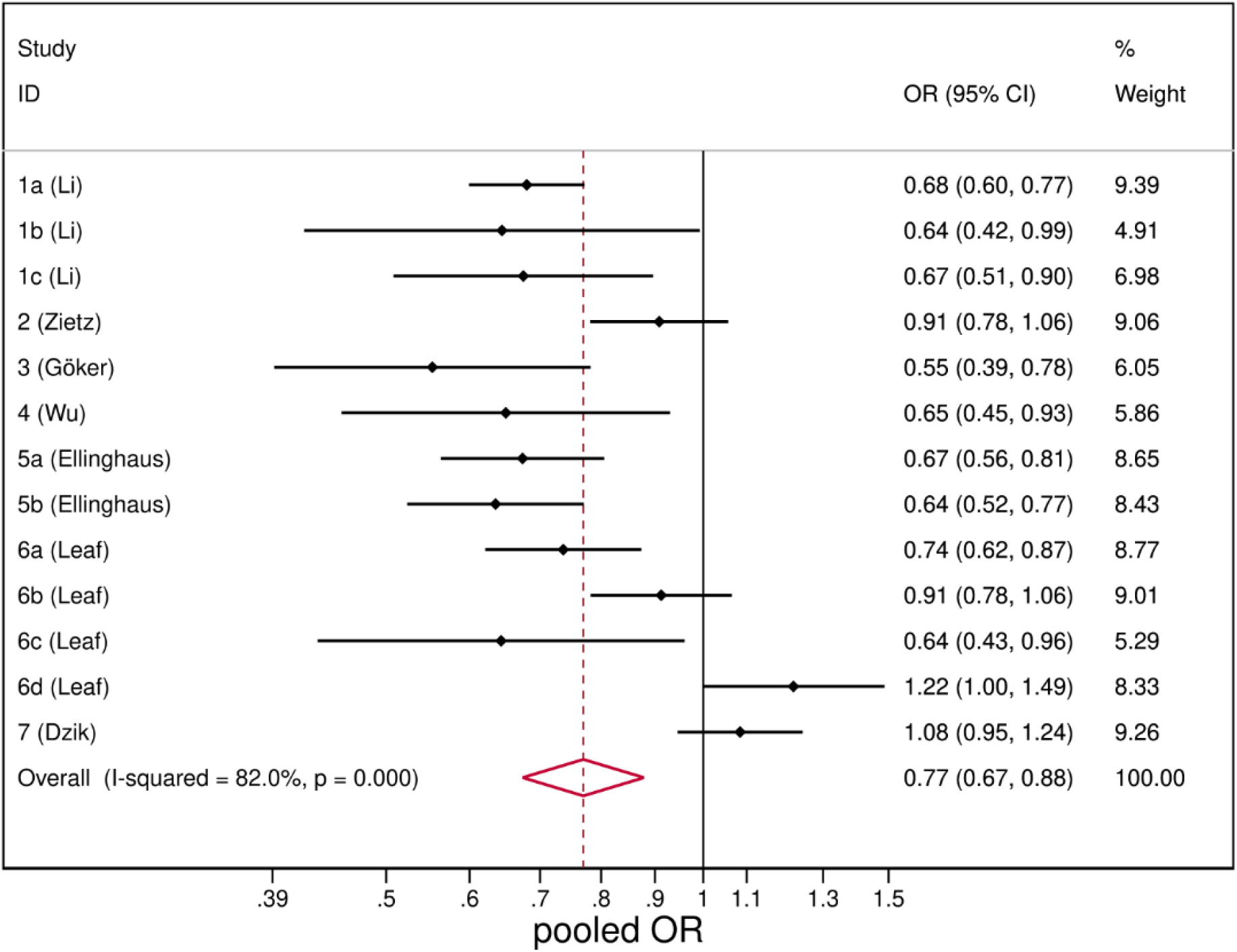
Forest plot from random effects analysis: OR of being blood group O in SARS-CoV-2+ group versus control group.

The type of control population did not explain the observed heterogeneity that remained considerable within each subgroup (Fig S3, Supplementary material). Again, grouping the studies by country we observed homogeneity among studies conducted in China (*I^2^*=0%) and the group of studies from Europe and Turkey (*I^2^*=0%), with pooled ORs significantly lower than the overall estimate (Fig S4, Supplementary material). Studies conducted in the USA instead reported contrasting results that led to a non-significant pooled estimate. The studies however have similar weights, thus the exclusion of one study in turn did not significantly affect the pooled estimate (Table S2, Supplementary material).

## Discussion

This pooled analysis included a large data sample extracted from available published studies on the association between ABO blood groups and SARS-CoV-2 infection. The findings of this meta-analysis suggest that blood group A is associated with an increase in the risk of SARS-CoV-2 infection, while in blood group O individuals the risk of being SARS-CoV-2+ is reduced.

Sensitivity and subgroup analysis showed similar results. Compared to controls, SARS-CoV-2+ patients appear to have blood group A more frequently and blood group O less frequently. We found no difference for blood groups B and AB. Results for blood group O are consistent for studies conducted in China, Europe and Turkey while there is much variability in the results of studies conducted in the USA. In particular, Leaf et al.^21^ differentiate the analysis for ethnicity, increasing the number of comparisons. It is within this study that much of the heterogeneity occurs. This heterogeneity could therefore be traced back to ethnicity, which should be taken into consideration - together with other possible related confounders - in designing future population studies to verify the influence of the ABO blood group on the kinetics of the pandemic, even within single countries. We found more homogeneity among the studies that use the general population as a control than among those that use blood donor populations or hospitalized patients.

Our analysis highlights the high heterogeneity in the type of studies and populations considered to date in the scientific literature on the topic. This is mainly due to the variability of settings and populations analysed and it is clearly an effect of the pandemic, which afflicted 188 countries around the world to date.

The presence of an association between genetic traits or specific blood groups is common for many diseases and several hypotheses have been formulated to explain it. Specifically, an increased host susceptibility associated with specific risk and predisposing factors in the host has been described for several infectious diseases.^5-9^ The ABO blood group polymorphism was seen to be associated in particular with 2002-2003 SARS-CoV infection. Many researchers argued whether this increased susceptibility is also present for SARS-CoV-2. Notably, several studies have reported the similarity of SARS-CoV-2 entry mechanism into target cells, exploiting the structural similarity of SARS-CoV and SARS-CoV-2 ACE1 and ACE2 receptors.^10-12^ The angiotensin I-converting enzyme (ACE1) and the more recently discovered homologue ACE2 are two antagonist enzymes of the RAS pathway that act and counterbalance each other.^11, 23-25^ The main role of ACE1 is the conversion of angiotensin I to angiotensin II, the latter being a peptide causing vasoconstriction, inflammation, fibrosis and proliferation. A high ACE2/ACE1 ratio protects against endothelial dysfunctions and vascular pathologies.^14,26^ SARS-CoV-2 enters human cells using the SARS-CoV receptor ACE2 and a specific transmembrane serine protease 2 (TMPRSS2) for the spike (S) protein priming.^11,12^ It has been described how a modest ACE2 expression characterizes the upper human respiratory tract and that this should limit the receptivity of the virus.^13,27^

Interestingly, a quantitative variation in ACE1 levels has been demonstrated to be modulated also by the ABO blood group locus. Moreover, selected ABO polymorphisms influence ACE inhibitors treatment response,^28-30^ and might contribute in reducing SARS-CoVs’ transmission, as reported by Guillon et al.,^9^ who ascribed to blood group O a lower risk of infection, hypothesizing that natural anti-A and anti-B antibodies can contribute in protecting against viral diseases at the population level.

Our analysis confirms an increased susceptibility, linked to ABO polymorphism, also for SARS-CoV-2. In particular, from our findings we can speculate that some risk factors in blood group A and protective characteristics in blood group O individuals might affect their biological response to the infection.

In order to expand our knowledge about the pathogen that is causing the most serious pandemic in modern human history, these insights are fundamental. The distribution of blood groups in the population and the awareness of their increased or reduced individual susceptibility to SARS-CoV-2 can be useful to understand the kinetics of the epidemic at the local level, and to implement population-level health policies and interventions aimed at reducing the viral spread.

This study has some limitations. First, we relied on the evidence and data available in the scientific literature to date, which are still preliminary. This is mainly due to the effect of the pandemic on research and publishing timing. This also influences the intrinsic quality of the studies included in the meta-analysis, which present a great variability in terms of study design and population considered. Second, we found it difficult to obtain a uniform adjustment for confounders. Third, the considerable size of the pooled control population that we reported is mainly due to data from a single study.^21^ Nevertheless, the size can still be deemed adequate for each study, when considered by itself. Lastly, both cases and controls populations may be deemed not adequately representative of the general population (e.g. most of included cases are hospitalized patients).

This article represents, to our knowledge, the first meta-analysis to investigate the epidemiological association of ABO blood group polymorphism with SARS-CoV-2 infection.

This meta-analysis provides additional evidence of the susceptibility of blood group A individuals to SARS-CoV-2 infection and the possible protective effect of blood group O.

Given the theoretical level of the underlying hypotheses, further methodologically sound studies are needed to investigate the molecular and clinical mechanism at the basis of the association between ABO polymorphism and individual susceptibility to SARS-CoV-2 infection.

## Data Availability

As a systematic-review and meta-analysis, this study uses data reported in other previously published articles, cited in the main text.

## Acknowledgments

None.

## Supporting informations

### Eligibility Criteria

#### Search strategy

**Table S1. Quality of studies assessment: Newcastle Ottawa Scale (NOS).**

**Table S2. Results from sensitivity analysis leave-one-out method: pooled OR and 95% confidence interval calculated omitting each study in turn.**

**Fig S1. Forest plot from random effects analysis: OR of being blood group A in SARS-CoV-2+ group versus control group, by type of control population.**

**Fig S2. Forest plot from random effects analysis: OR of being blood group A in SARS-CoV-2+ group versus control group, by Country.**

**Fig S3. Forest plot from random effects analysis: OR of being blood group 0 in SARS-CoV-2+ group versus control group, by type of control population.**

**Fig S4. Forest plot from random effects analysis: OR of being blood group 0 in SARS-CoV-2+ group versus control group, by Country.**

